# Improving Clinical Applicability of Heart Failure Readmission Prediction via Automated Feature Engineering

**DOI:** 10.64898/2026.02.26.26346970

**Authors:** M. Oloko-Oba, A. Aslam, M. Echols, A. Onwuanyi, M.Y. Idris

## Abstract

Heart failure (HF) readmission prediction models often rely on manually curated, cross-sectional features and show limited discrimination and calibration. We evaluated whether automated feature engineering via Deep Feature Synthesis (DFS) improves the clinical applicability of HF readmission prediction from longitudinal electronic health record data. Using 355,217 HF hospitalizations from a large U.S. safety-net health system (2010–2025), we compared a clinician-curated baseline feature set to DFS-enhanced features and trained identical models for 30-, 60-, and 90-day readmission. DFS consistently improved gradient-boosted tree performance, increasing AUROC and AUPRC across all horizons, while logistic regression performance declined. At sensitivity-targeted operating points (80%), DFS improved specificity and positive predictive value for boosted trees, reducing false-positive workload. Calibration also improved for boosted trees at all horizons but not for linear models. These results show that automated feature engineering yields deployment-relevant gains that are strongly model-class dependent.

**Data and Code Availability:** This study uses retrospective electronic health record data from a large urban safety-net healthcare system in the United States. Due to patient privacy, institutional restrictions, and data use agreements, the data are not publicly available. An anonymized version of the code used for data processing, feature engineering, model training, and evaluation will be made available upon acceptance of the paper.

**Institutional Review Board (IRB):** This retrospective study was reviewed and approved by an institutional review board. Full IRB details will be provided in the camera-ready version of the paper if accepted.

## 1. Introduction

Heart failure (HF) remains a leading cause of hospitalization worldwide, affecting roughly 64 million people and occurring in 1-3% of adults (Fine et al., 2024; Yu and Son, 2024). HF readmission remains a major clinical concern among patients, with an estimated 20%–25% returning within 30 days of discharge and even higher rates by 90 days, despite widespread implementation of guideline-directed care and disease-management programs (Abdul-Samad et al., 2024; Martin et al., 2024). These early readmissions not only impose substantial financial burden but also signal potentially preventable adverse outcomes and disruptions in continuity of care (Yu and Son, 2024). Accurately identifying patients at high risk of 30-, 60-, or 90-day readmission is a vital clinical and policy priority.

Over the past two decades, numerous clinical prediction models have been developed to predict HF readmission using conventional techniques applied to relatively small demographic characteristics, comorbidities, length of stay, vital signs, and laboratory values that are manually engineered (Fine et al., 2024; Martin et al., 2024; Van Grootven et al., 2021; Rahimi et al., 2014). These approaches often overlook the rich temporal information (e.g., trajectories of vitals and labs, patterns of prior utilization) and complex interactions that are embedded in high-dimensional electronic health records (EHR) data. As a result, most of these models continue to achieve only modest discrimination and poor calibration (Van Grootven et al., 2021; Ouwerkerk et al., 2014; Frizzell et al., 2017).

One reason many existing heart failure readmission models achieve limited clinical applicability is their reliance on manually curated, cross-sectional features (Davis et al., 2022). Representation learning approaches for heart failure risk prediction can exploit longitudinal EHR data but typically rely on large integrated datasets and require substantial data, tuning, and infrastructure (Golas et al., 2018). This motivates interest in feature-based methods that can systematically incorporate the temporal and relational structure of clinical encounters while remaining interpretable and suitable for clinically calibrated risk prediction.

Automated feature engineering offers an opportunity to systematically expand clinically grounded feature spaces by transforming expert-identified variables into longitudinal and relational representations (Luo, 2019; Alnegheimish et al., 2020). By applying consistent aggregation and transformation operations across encounters, automated approaches can expose temporal patterns, recurrence, and interactions that are difficult to enumerate manually, while remaining anchored in clinically meaningful inputs. In this way, automated feature engineering can serve as a structured mechanism to augment clinician-curated features derived from EHR data.

In this study, we investigate whether automated feature engineering via Deep Feature Synthesis (DFS) improves HF readmission prediction from longitudinal EHR data compared to a clinician-curated baseline feature set. We build upon prior applications of DFS in heart failure risk prediction Fine et al. (2024) by extending evaluation beyond single time horizons and standard discrimination metrics. We benchmark performance across multiple model families, including logistic regression, gradient-boosted decision trees (LightGBM/XGBoost), and a multilayer perceptron (MLP), at 30-, 60-, and 90-day horizons, emphasizing not only discrimination but also calibration and operating characteristics at clinically relevant sensitivity targets. Specifically, we ask: (1) whether DFS-enhanced features improve discrimination and calibration over the baseline across time horizons; (2) whether improvements are consistent across model classes; and (3) how DFS affects threshold-dependent operating performance.

## 2. Methods

We conducted a retrospective prediction modeling study using electronic health record data to estimate 30-, 60-, and 90-day readmission risk following heart failure hospitalization. The study is reported in accordance with the TRIPOD+AI reporting guideline.

### 2.1. Study Population and Data Sources

We assembled an encounter-level cohort from a large urban safety-net healthcare system in the United States using electronic health record (EHR) data extracted from a commercial clinical information system. The dataset spanned February 2010 through April 2025 and included patient demographics, encounter-level admission and discharge information, diagnoses and procedures, laboratory results, vital signs, and medication data (orders, prescriptions, and administrations) recorded as part of routine clinical care.

The analytic cohort comprised 355,217 eligible heart failure–related index hospitalizations (encounter-level observations). Baseline characteristics summarized in Table 1 include age at admission, sex, race/ethnicity, body mass index, smoking status, and routinely recorded vital signs (blood pressure, heart rate, respiratory rate, and temperature). Age was reported for admissions with valid adult values (18–110 years), treating age values of 0 as invalid. Observed readmission rates were 15.8% at 30 days, 22.0% at 60 days, and 25.9% at 90 days.

**Table 1:** Demographics and readmission incidence.

| Characteristic | Total (n = 355,217) |
| --- | --- |
| Age at admission, years | 52.66 ± 17.66 |
| **Sex** |  |
| Female | 175,129 (49.3%) |
| Male | 180,017 (50.7%) |
| **Race** |  |
| American Indian or Alaska Native | 1,053 (0.3%) |
| Asian | 3,276 (0.9%) |
| Black or African American | 285,786 (80.5%) |
| Native Hawaiian or Other Pacific Islander | 263 (0.1%) |
| White or Caucasian | 39,671 (11.2%) |
| Other / Unknown / Refused<sup>†</sup> | 25,508 (7.2%) |
| **Ethnicity** |  |
| Hispanic or Latino | 21,752 (6.1%) |
| Not Hispanic or Latino | 327,319 (92.1%) |
| Unknown ethnicity | 5,159 (1.5%) |
| **Readmission incidence** |  |
| 30-day readmission | 56,094 (15.8%) |
| 60-day readmission | 78,211 (22.0%) |
| 90-day readmission | 92,124 (25.9%) |

<sup>†</sup>Other / Unknown / Refused includes records labeled as other race, other, patient refused, unknown, or unspecified.

To minimize information leakage, cohort assembly and feature generation were anchored to the index hospitalization and restricted to data available up to discharge (or the prespecified prediction time point), and outcomes were assessed only after that point.

### 2.2. Outcomes

The primary outcome was 30-day readmission, defined as any inpatient readmission occurring within 30 days after discharge from the index hospitalization. Secondary outcomes were 60-day and 90-day readmission, defined analogously. Modeling each horizon separately allowed us to assess whether automated feature engineering provides consistent value across short-term transitional care decisions and longer post-discharge vulnerability windows.

We designated 30-day readmission as the primary outcome due to its clinical and policy relevance, while treating 60- and 90-day readmission as secondary outcomes to assess whether automated feature engineering provides greater benefit beyond the immediate post-discharge period.

### 2.3. Features

We compared two feature construction strategies, each yielding a distinct feature set:

1. *Clinical-Curated (Baseline):* The clinician-curated baseline feature construction was developed through an iterative, collaborative process involving three practicing cardiologists. Across multiple rounds, cardiologists proposed candidate predictors based on their clinical expertise and experience managing heart failure, reviewed intermediate feature representations, and refined selections through discussion to arrive at a consensus feature set drawn from commonly used EHR domains, including demographics and routinely summarized clinical measures. As part of this process, selected raw EHR fields with non-informative feature names (e.g., generic medication fields) were manually normalized into clinically meaningful variables, such as average administered dose per encounter. This workflow reflects how expert clinical judgment is translated into operational risk-stratification pipelines, prioritizing clinical relevance, interpretability, and feasibility for real-world deployment.
2. *Automated Feature Construction (Deep Feature Synthesis):* Automated feature construction was implemented using Deep Feature Synthesis (DFS) applied to encounter-level, multidomain EHR table, including laboratory results, vital signs, medication administrations and prescriptions, and procedures associated with each hospitalization. DFS was configured with shallow depth (maximum depth = 1) and a predefined primitive set. Aggregation primitives included mean, maximum, minimum, and count, and datetime transformations included month, weekday, hour, and weekend indicators. Feature generation was performed in batches (chunk size = 20,000) with a cap of 5,000 total features, using table-specific ignore lists to exclude non-informative or high-risk columns. To prevent temporal leakage, all features were generated using discharge time as the encounter-specific cut-off, restricting inputs to information available prior to discharge.

The clinician-curated baseline feature set comprised approximately XX features, whereas DFS generated up to 5,000 candidate features per horizon, of which Y features were retained after preprocessing and filtering.

### 2.4. Model Development

#### 2.4.1. Missing Data

Missingness varied substantially across candidate predictors and was not consistent with a missing-completely-at-random (MCAR) assumption, based on feature-level missingness rates and patterns of co-missingness across related variables. We employed a hybrid, computationally tractable strategy. Predictors with extreme missingness (*>*80%) and near-zero variance were excluded. A small, prespecified subset of clinically central demographic and clinical variables (20 features) was imputed using MICEforest^1^, while all remaining variables were imputed using single imputation (median for numeric variables; mode for categorical variables). Missingness indicators were retained where informative to capture potential missing-not-at-random patterns.

#### 2.4.2. Model Selection

To isolate the effect of feature engineering independent of model choice, we trained the same set of model families on both the baseline and DFS-enhanced feature sets for each outcome horizon. Models included logistic regression (LR) as a transparent linear baseline, gradient-boosted decision trees (LightGBM and XGBoost) to capture nonlinear interactions in structured data, and a multilayer perceptron (MLP) as a neural baseline for nonlinear function approximation. Separate models were trained for 30-, 60-, and 90-day readmission.

### 2.5. Validation Strategy

Model evaluation used held-out data partitions designed to reflect prospective deployment. Splitting was performed to prevent patient-level leakage across training, validation, and test sets by assigning all encounters from a given patient to a single partition. Each hospitalization was treated as an independent index event, with patient-level partitioning used to ensure that all encounters from a given patient were assigned to a single data split. Hyperparameters were tuned using the validation set only, optimizing AU-ROC with identical search spaces across baseline and DFS-enhanced feature sets. Splits were fixed across all model comparisons.

### 2.5. Performance Evaluation

Because hospital readmission prediction is a class-imbalanced problem, we reported complementary performance metrics capturing different aspects of model behavior. Discrimination was assessed using the area under the receiver operating characteristic curve (AUROC), summarizing ranking ability across thresholds. Performance under class imbalance was assessed using the area under the precision–recall curve (AUPRC), which is often more informative than AUROC when outcomes are relatively infrequent. In addition, we reported threshold-based metrics including sensitivity, specificity, positive predictive value (PPV), and negative predictive value (NPV), each with 95% confidence intervals. Confidence intervals for all threshold-based and discrimination metrics were estimated using nonparametric bootstrapping on the held-out test set.

We also generated receiver operating characteristic (ROC) and precision–recall (PR) curves for each prediction horizon, overlaying baseline and DFS-enhanced models within each model family to examine how performance differs across operating points beyond single-metric summaries.

#### 2.6.1. Operating Points AND Threshold Selection

To support clinical interpretability, threshold-based performance was evaluated in two ways. First, we reported metrics at a fixed probability threshold of 0.50, providing a simple and reproducible reference point for baseline comparisons. Second, because many readmission-reduction workflows prioritize identifying high-risk patients, we selected decision thresholds to achieve prespecified sensitivity targets (i.e., 0.80) and reported the corresponding specificity, PPV, and NPV. This framing reflects how risk scores are often operationalized in practice, where a desired capture rate is specified and the associated false-positive burden is then evaluated.

### 2.7. Calibration Assessment

Model calibration was assessed using the Brier score, a proper scoring rule that measures the mean squared error of predicted probabilities, with lower values indicating better calibration. Brier scores were computed on the held-out test set for each model family and prediction horizon (30, 60, and 90 days). To contextualize absolute performance, we also report the Brier Skill Score (BSS), defined relative to a null model that predicts the observed event prevalence; positive BSS values indicate improvement over the null benchmark.Differences in calibration between baseline and DFS-enhanced models were evaluated using paired comparisons on the same test partitions. Statistical significance was assessed using nonparametric bootstrap confidence intervals for paired differences in test-set Brier scores, quantifying whether observed calibration differences exceeded what would be expected from sampling variability alone.

## 3. Results

### 3.1. Cohort and 30/60/90 readmission rates

The final analytic cohort included 355,217 eligible heart failure–related index hospitalizations. The mean age at admission was 52.66 ± 17.66 years; 49.3% of encounters were among female patients, and the cohort was predominantly Black or African American (80.5%). Readmission incidence increased with the prediction horizon, occurring in 15.8% of encounters within 30 days, 22.0% within 60 days, and 25.9% within 90 days of discharge. Detailed demographic and baseline clinical characteristics are reported in Table 1.^2^

### 3.2. Discrimination and precision–recall by horizon and model family

Table 2 summarizes discrimination (AUROC) and precision–recall performance (AUPRC) for clinician-curated baseline LR and LightGBM models and DFS-enhanced models at 30-, 60-, and 90-day horizons. To isolate the contribution of automated feature engineering, we focus on within-model deltas (DFS − baseline) rather than absolute values.

**Table 2:** Discrimination (AUROC) and precision–recall performance (AUPRC) for clinician-curated baseline and DFS-enhanced Logistic Regression and LightGBM models across prediction horizons.

| Horizon | Model | AUROC (Base / DFS) | ΔAUROC | AUPRC (Base / DFS) | ΔAUPRC |
| --- | --- | --- | --- | --- | --- |
| 2*30-day | LR | 0.704 / 0.691 | −0.013 | 0.325 / 0.313 | −0.012 |
|  | LightGBM | 0.712 / 0.728 | +0.016 | 0.348 / 0.367 | +0.019 |
| 2*60-day | LR | 0.712 / 0.697 | −0.015 | 0.415 / 0.396 | −0.019 |
|  | LightGBM | 0.718 / 0.733 | +0.015 | 0.435 / 0.453 | +0.018 |
| 2*90-day | LR | 0.710 / 0.695 | −0.015 | 0.461 / 0.441 | −0.020 |
|  | LightGBM | 0.723 / 0.739 | +0.016 | 0.485 / 0.506 | +0.021 |

Deltas ( $\Delta$ ) indicate DFS minus baseline performance on the held-out test set.

LightGBM consistently achieved the strongest overall performance across discrimination, operating characteristics, and calibration. Accordingly, we focus the main results on comparisons between logistic regression (as a clinically interpretable baseline) and LightGBM (as the best-performing model), with additional model families reported in the Supplementary Appendix.

For LightGBM, DFS produced consistent improvements in both AUROC and AUPRC across all horizons. AUROC increased by +0.016 at 30 days, +0.015 at 60 days, and +0.016 at 90 days. Corresponding gains in AUPRC were +0.019, +0.018, and +0.021, respectively. These improvements were consistent in direction and magnitude across time horizons. In contrast, logistic regression showed neutral or negative changes with DFS. AUROC decreased by − 0.013 at 30 days, − 0.015 at 60 days, and − 0.015 at 90 days, with corresponding declines in AUPRC of − 0.012, − 0.019, and − 0.020. Across horizons, DFS did not improve discrimination or precision for logistic regression.

### 3.3. Operating characteristics

#### 3.3.1. Fixed probability threshold (0.50)

At a fixed threshold of 0.50 fixed threshold, logistic regression models consistently favored sensitivity over precision, identifying a larger fraction of true readmissions at the expense of lower positive predictive value (PPV). For example, at 30 days, the baseline logistic regression model achieved a sensitivity of 0.625 with a PPV of 0.265, indicating broad case capture but substantial false-positive burden.

In contrast, LightGBM models exhibited the opposite operating profile, with substantially higher specificity and PPV but lower sensitivity. The DFS-enhanced LightGBM model at 30 days achieved a specificity of 0.896 and PPV of 0.391, while sensitivity decreased to 0.341. Similar patterns were observed at 60- and 90-day horizons, reflecting a consistent tradeoff between precision and case capture across time windows.

### 3.3.2. Sensitivity-targeted operating points

Table 3 summarizes model performance at sensitivity-targeted operating points, where decision thresholds were selected to achieve a prespecified sensitivity of 0.80 and the resulting specificity and predictive values were examined. This evaluation reflects deployment scenarios in which health systems prioritize identifying most patients at risk for readmission while managing false-positive burden.

**Table 3:** Performance of readmission prediction models at operating points targeting 80% sensitivity for 30-, 60-, and 90-day readmission.

| Horizon | Model | Sensitivity | Specificity | PPV | NPV |
| --- | --- | --- | --- | --- | --- |
| 4*30-day | LR_Base | 0.800 (0.792–0.809) | 0.468 (0.463–0.473) | 0.228 (0.223–0.232) | 0.923 (0.920–0.926) |
|  | LR_DFS | 0.800 (0.792–0.809) | 0.444 (0.440–0.449) | 0.220 (0.215–0.224) | 0.919 (0.916–0.923) |
|  | LGBM_Base | 0.800 (0.792–0.808) | 0.470 (0.465–0.474) | 0.228 (0.223–0.233) | 0.923 (0.920–0.927) |
|  | LGBM_DFS | 0.800 (0.792–0.809) | 0.502 (0.497–0.507) | 0.239 (0.234–0.244) | 0.928 (0.925–0.931) |
| 4*60-day | LR_Base | 0.800 (0.792–0.807) | 0.491 (0.486–0.496) | 0.318 (0.313–0.323) | 0.892 (0.888–0.896) |
|  | LR_DFS | 0.800 (0.793–0.808) | 0.468 (0.463–0.473) | 0.309 (0.304–0.314) | 0.888 (0.884–0.892) |
|  | LGBM_Base | 0.800 (0.792–0.807) | 0.490 (0.485–0.495) | 0.317 (0.312–0.322) | 0.892 (0.888–0.896) |
|  | LGBM_DFS | 0.800 (0.793–0.807) | 0.514 (0.509–0.519) | 0.328 (0.323–0.333) | 0.897 (0.893–0.900) |
| 4*90-day | LR_Base | 0.800 (0.794–0.807) | 0.486 (0.481–0.491) | 0.363 (0.358–0.369) | 0.869 (0.865–0.874) |
|  | LR_DFS | 0.800 (0.794–0.807) | 0.462 (0.458–0.467) | 0.353 (0.348–0.358) | 0.863 (0.859–0.868) |
|  | LGBM_Base | 0.800 (0.793–0.806) | 0.500 (0.495–0.505) | 0.369 (0.364–0.375) | 0.872 (0.868–0.876) |
|  | LGBM_DFS | 0.800 (0.793–0.807) | 0.526 (0.521–0.531) | 0.382 (0.377–0.388) | 0.878 (0.874–0.882) |

Values are reported as estimate (95% confidence interval). Model abbreviations: LR\_Base = logistic regression baseline; LR\_DFS = logistic regression with DFS; LGBM\_Base = LightGBM baseline; LGBM\_DFS = LightGBM with DFS.

Logistic regression showed reduced specificity and PPV after DFS at matched sensitivity, despite identical case capture. At 30 days, baseline logistic regression achieved a specificity of 0.468 and PPV of 0.228, whereas the DFS-enhanced model achieved a specificity of 0.444 and PPV of 0.220. This pattern persisted across longer horizons.

At matched sensitivity, DFS-enhanced LightGBM consistently improved specificity and positive predictive value (PPV) across all horizons (Table 3). At 30 days, DFS increased specificity from 0.470 to 0.502 (+0.032) and PPV from 0.228 to 0.239 (+0.011) relative to baseline LightGBM. Similar improvements were observed at 60 days (specificity +0.024; PPV +0.011) and 90 days (specificity +0.027; PPV +0.013).

These gains translate into meaningful reductions in false-positive workload. For example, at 30 days, a PPV of 0.239 corresponds to approximately 4.18 alerts per true readmission, compared with 4.39 alerts per true readmission for baseline LightGBM at the same sensitivity. Comparable reductions were observed at 60 and 90 days.

### 3.4. Calibration

Table 4 summarizes calibration performance across model families and prediction horizons. For Light-GBM, DFS consistently improved calibration at all horizons, yielding lower Brier scores with paired differences (Brier = DFS baseline) whose 95% bootstrap confidence intervals did not cross zero. At 30 days, DFS reduced the Brier score from − 0.1596 to − 0.1572 (Brier − 0.0025, 95% CI − 0.0029 to − 0.0020). Similar improvements were observed at 60 days (Brier − 0.0027, 95% CI − 0.0032 to − 0.0022) and 90 days (Brier − 0.0035, 95% CI − 0.0039 to − 0.0030). These reductions corresponded to increased Brier Skill Scores at 60 and 90 days, indicating improved calibration relative to the null model.

**Table 4:** Test-set (Brier) calibration performance for baseline and DFS-enhanced Logistic Regression and LightGBM models across prediction horizons.

| Horizon | Model | Brier (Base) | Brier (DFS) | BSS (Base / DFS) | ΔBrier (95% CI) |
| --- | --- | --- | --- | --- | --- |
| 30-day | LR | 0.2159 | 0.2165 | −0.577 / −0.582 | +0.0006 (−0.0003, +0.0015) |
| 30-day | LGBM | 0.1596 | 0.1572 | −0.166 / −0.148 | −0.0025 (−0.0029, −0.0020) |
| 60-day | LR | 0.2161 | 0.2159 | −0.225 / −0.224 | −0.0001 (−0.0009, +0.0006) |
| 60-day | LGBM | 0.1729 | 0.1701 | +0.020 / +0.035 | −0.0027 (−0.0032, −0.0022) |
| 90-day | LR | 0.2159 | 0.2202 | −0.100 / −0.122 | +0.0043 (+0.0034, +0.0051) |
| 90-day | LGBM | 0.1800 | 0.1765 | +0.083 / +0.101 | −0.0035 (−0.0039, −0.0030) |

Confidence intervals were estimated using paired stratified bootstrapping.

**Table 5:** Demographic and baseline clinical characteristics of the study cohort.

| **Characteristic** | **Total (n = 355,217)** |
| --- | --- |
| Age at admission, years | 52.66 ± 17.66 |
| **Sex** |  |
| Female | 175,129 (49.3%) |
| Male | 180,017 (50.7%) |
| **Race** |  |
| American Indian or Alaska Native | 1,053 (0.3%) |
| Asian | 3,276 (0.9%) |
| Black or African American | 285,786 (80.5%) |
| Native Hawaiian or Other Pacific Islander | 263 (0.1%) |
| White or Caucasian | 39,671 (11.2%) |
| Other / Unknown / Refused<sup>†</sup> | 25,508 (7.2%) |
| **Ethnicity** |  |
| Hispanic or Latino | 21,752 (6.1%) |
| Not Hispanic or Latino | 327,319 (92.1%) |
| Unknown ethnicity | 5,159 (1.5%) |
| **Vital signs** |  |
| Body mass index, kg/m<sup>2</sup> | 27.68 ± 7.42 |
| Systolic blood pressure, mmHg | 130.24 ± 18.87 |
| Diastolic blood pressure, mmHg | 76.20 ± 11.92 |
| Heart rate, beats/min | 80.79 ± 9.95 |
| Respiratory rate, breaths/min | 18.17 ± 1.97 |
| Temperature, °F | 98.12 ± 0.52 |
| **Smoking status** |  |
| Never smoker | 117,576 (33.1%) |
| Former smoker | 61,902 (17.4%) |
| Current smoker | 87,348 (24.6%) |
| Light smoker | 1,977 (0.6%) |
| Heavy smoker | 561 (0.2%) |
| Unknown smoking status | 85,452 (24.1%) |
| **Readmission incidence** |  |
| 30-day readmission | 56,094 (15.8%) |
| 60-day readmission | 78,211 (22.0%) |
| 90-day readmission | 92,124 (25.9%) |

<sup>†</sup>Other / Unknown / Refused includes records labeled as other race, other, patient refused, unknown, or unspecified.

**Table 6:** Detailed performance of all prediction models at a fixed probability threshold of 0.50 for 30-, 60-, and 90-day readmission. Values are reported as estimate (95% confidence interval).

| Horizon | Model | Sensitivity | Specificity | PPV | NPV | AUROC | AUPRC |
| --- | --- | --- | --- | --- | --- | --- | --- |
| 8*30-day | LR_Base | 0.625 (0.615–0.636) | 0.660 (0.656–0.665) | 0.265 (0.260–0.270) | 0.900 (0.896–0.903) | 0.704 (0.698–0.710) | 0.325 (0.317–0.333) |
|  | LR_DFS | 0.567 (0.557–0.577) | 0.704 (0.700–0.708) | 0.273 (0.266–0.279) | 0.893 (0.889–0.896) | 0.691 (0.686–0.697) | 0.313 (0.306–0.322) |
|  | LGBM_Base | 0.279 (0.269–0.288) | 0.914 (0.911–0.916) | 0.388 (0.375–0.400) | 0.866 (0.863–0.869) | 0.712 (0.706–0.717) | 0.348 (0.339–0.356) |
|  | LGBM_DFS | 0.341 (0.331–0.351) | 0.896 (0.893–0.899) | 0.391 (0.380–0.402) | 0.874 (0.871–0.877) | 0.728 (0.722–0.733) | 0.367 (0.358–0.376) |
|  | XGB_Base | 0.219 (0.211–0.228) | 0.937 (0.934–0.939) | 0.403 (0.390–0.417) | 0.860 (0.857–0.863) | 0.702 (0.697–0.708) | 0.339 (0.331–0.348) |
|  | XGB_DFS | 0.285 (0.276–0.295) | 0.917 (0.914–0.919) | 0.401 (0.389–0.414) | 0.868 (0.865–0.871) | 0.717 (0.712–0.723) | 0.356 (0.347–0.366) |
|  | MLP_Base | 0.160 (0.153–0.168) | 0.934 (0.931–0.936) | 0.321 (0.307–0.335) | 0.850 (0.847–0.853) | 0.654 (0.648–0.660) | 0.261 (0.255–0.268) |
|  | MLP_DFS | 0.048 (0.044–0.053) | 0.981 (0.980–0.982) | 0.334 (0.308–0.360) | 0.840 (0.837–0.844) | 0.630 (0.624–0.636) | 0.241 (0.236–0.247) |
| 8*60-day | LR_Base | 0.655 (0.647–0.664) | 0.646 (0.641–0.650) | 0.354 (0.348–0.360) | 0.863 (0.859–0.867) | 0.712 (0.706–0.716) | 0.415 (0.407–0.423) |
|  | LR_DFS | 0.597 (0.589–0.606) | 0.676 (0.671–0.680) | 0.353 (0.346–0.360) | 0.850 (0.846–0.854) | 0.697 (0.692–0.702) | 0.396 (0.389–0.403) |
|  | LGBM_Base | 0.313 (0.305–0.321) | 0.898 (0.895–0.901) | 0.477 (0.465–0.488) | 0.815 (0.811–0.819) | 0.718 (0.714–0.724) | 0.435 (0.427–0.443) |
|  | LGBM_DFS | 0.359 (0.351–0.368) | 0.886 (0.882–0.889) | 0.482 (0.471–0.492) | 0.823 (0.819–0.827) | 0.733 (0.729–0.738) | 0.453 (0.446–0.461) |
|  | XGB_Base | 0.227 (0.220–0.234) | 0.932 (0.929–0.934) | 0.495 (0.482–0.508) | 0.803 (0.799–0.806) | 0.703 (0.698–0.708) | 0.420 (0.413–0.428) |
|  | XGB_DFS | 0.296 (0.288–0.304) | 0.910 (0.907–0.913) | 0.493 (0.482–0.505) | 0.813 (0.810–0.817) | 0.723 (0.718–0.728) | 0.442 (0.435–0.450) |
|  | MLP_Base | 0.150 (0.144–0.156) | 0.938 (0.935–0.940) | 0.417 (0.402–0.431) | 0.788 (0.785–0.792) | 0.652 (0.647–0.658) | 0.339 (0.333–0.346) |
|  | MLP_DFS | 0.137 (0.131–0.143) | 0.936 (0.934–0.939) | 0.390 (0.375–0.404) | 0.785 (0.782–0.789) | 0.640 (0.634–0.645) | 0.327 (0.321–0.333) |
| 8*90-day | LR_Base | 0.662 (0.654–0.670) | 0.636 (0.631–0.641) | 0.400 (0.393–0.406) | 0.837 (0.832–0.841) | 0.710 (0.705–0.715) | 0.461 (0.453–0.468) |
|  | LR_DFS | 0.593 (0.585–0.601) | 0.678 (0.673–0.683) | 0.403 (0.396–0.410) | 0.820 (0.815–0.824) | 0.695 (0.690–0.700) | 0.441 (0.434–0.448) |
|  | LGBM_Base | 0.315 (0.308–0.323) | 0.895 (0.892–0.898) | 0.524 (0.513–0.535) | 0.781 (0.777–0.785) | 0.723 (0.718–0.728) | 0.485 (0.478–0.492) |
|  | LGBM_DFS | 0.370 (0.363–0.378) | 0.882 (0.879–0.885) | 0.535 (0.525–0.545) | 0.793 (0.788–0.796) | 0.739 (0.735–0.744) | 0.506 (0.498–0.513) |
|  | XGB_Base | 0.236 (0.229–0.243) | 0.929 (0.927–0.932) | 0.550 (0.537–0.562) | 0.769 (0.765–0.772) | 0.708 (0.703–0.713) | 0.469 (0.462–0.476) |
|  | XGB_DFS | 0.297 (0.290–0.305) | 0.910 (0.907–0.912) | 0.546 (0.535–0.557) | 0.779 (0.776–0.783) | 0.728 (0.723–0.732) | 0.492 (0.485–0.500) |
|  | MLP_Base | 0.249 (0.242–0.256) | 0.898 (0.894–0.900) | 0.471 (0.460–0.482) | 0.765 (0.762–0.769) | 0.668 (0.663–0.673) | 0.414 (0.407–0.421) |
|  | MLP_DFS | 0.165 (0.159–0.172) | 0.917 (0.914–0.920) | 0.422 (0.410–0.435) | 0.750 (0.746–0.754) | 0.644 (0.639–0.649) | 0.371 (0.365–0.377) |

**Table 7:** Full performance metrics for all prediction models at operating points targeting 80% sensitivity for 30-, 60-, and 90-day readmission. Values are reported as estimate (95% confidence interval).

| Horizon | Model | Threshold | Sensitivity | Specificity | PPV | NPV |
| --- | --- | --- | --- | --- | --- | --- |
| 8*30-day | LR_Base | 0.4265 | 0.800 (0.792–0.809) | 0.468 (0.463–0.473) | 0.228 (0.223–0.232) | 0.923 (0.920–0.926) |
|  | LR_DFS | 0.3530 | 0.800 (0.792–0.809) | 0.444 (0.440–0.449) | 0.220 (0.215–0.224) | 0.919 (0.916–0.923) |
|  | LGBM_Base | 0.3205 | 0.800 (0.792–0.808) | 0.470 (0.465–0.474) | 0.228 (0.223–0.233) | 0.923 (0.920–0.927) |
|  | LGBM_DFS | 0.3195 | 0.800 (0.792–0.809) | 0.502 (0.497–0.507) | 0.239 (0.234–0.244) | 0.928 (0.925–0.931) |
|  | XGB_Base | 0.3270 | 0.801 (0.793–0.809) | 0.452 (0.448–0.457) | 0.223 (0.218–0.227) | 0.921 (0.917–0.924) |
|  | XGB_DFS | 0.3225 | 0.800 (0.792–0.808) | 0.483 (0.479–0.488) | 0.233 (0.228–0.237) | 0.925 (0.922–0.928) |
|  | MLP_Base | 0.3090 | 0.800 (0.791–0.808) | 0.397 (0.393–0.402) | 0.206 (0.202–0.210) | 0.910 (0.906–0.914) |
|  | MLP_DFS | 0.3235 | 0.801 (0.793–0.809) | 0.360 (0.355–0.364) | 0.197 (0.193–0.201) | 0.902 (0.898–0.907) |
| 8*60-day | LR_Base | 0.4370 | 0.800 (0.792–0.807) | 0.491 (0.486–0.496) | 0.318 (0.313–0.323) | 0.892 (0.888–0.896) |
|  | LR_DFS | 0.4030 | 0.800 (0.793–0.808) | 0.468 (0.463–0.473) | 0.309 (0.304–0.314) | 0.888 (0.884–0.892) |
|  | LGBM_Base | 0.3310 | 0.800 (0.792–0.807) | 0.490 (0.485–0.495) | 0.317 (0.312–0.322) | 0.892 (0.888–0.896) |
|  | LGBM_DFS | 0.3280 | 0.800 (0.793–0.807) | 0.514 (0.509–0.519) | 0.328 (0.323–0.333) | 0.897 (0.893–0.900) |
|  | XGB_Base | 0.3335 | 0.801 (0.794–0.808) | 0.460 (0.455–0.465) | 0.305 (0.300–0.310) | 0.886 (0.882–0.890) |
|  | XGB_DFS | 0.3315 | 0.799 (0.792–0.806) | 0.499 (0.494–0.504) | 0.321 (0.316–0.326) | 0.893 (0.889–0.897) |
|  | MLP_Base | 0.3010 | 0.799 (0.792–0.806) | 0.403 (0.398–0.408) | 0.284 (0.279–0.289) | 0.871 (0.867–0.876) |
|  | MLP_DFS | 0.3670 | 0.800 (0.793–0.807) | 0.381 (0.376–0.385) | 0.277 (0.272–0.282) | 0.865 (0.860–0.870) |
| 8*90-day | LR_Base | 0.4380 | 0.800 (0.794–0.807) | 0.486 (0.481–0.491) | 0.363 (0.358–0.369) | 0.869 (0.865–0.874) |
|  | LR_DFS | 0.4055 | 0.800 (0.794–0.807) | 0.462 (0.458–0.467) | 0.353 (0.348–0.358) | 0.863 (0.859–0.868) |
|  | LGBM_Base | 0.3365 | 0.800 (0.793–0.806) | 0.500 (0.495–0.505) | 0.369 (0.364–0.375) | 0.872 (0.868–0.876) |
|  | LGBM_DFS | 0.3345 | 0.800 (0.793–0.807) | 0.526 (0.521–0.531) | 0.382 (0.377–0.388) | 0.878 (0.874–0.882) |
|  | XGB_Base | 0.3390 | 0.800 (0.793–0.806) | 0.470 (0.466–0.475) | 0.356 (0.351–0.361) | 0.865 (0.861–0.870) |
|  | XGB_DFS | 0.3380 | 0.800 (0.793–0.806) | 0.509 (0.504–0.514) | 0.374 (0.368–0.379) | 0.874 (0.870–0.878) |
|  | MLP_Base | 0.3225 | 0.800 (0.794–0.807) | 0.415 (0.410–0.419) | 0.334 (0.329–0.339) | 0.850 (0.845–0.855) |
|  | MLP_DFS | 0.3585 | 0.799 (0.793–0.806) | 0.392 (0.388–0.397) | 0.325 (0.320–0.330) | 0.842 (0.837–0.847) |

**Table 8:** Test-set calibration relative to the null benchmark for all model families across 30-, 60-, and 90-day readmission horizons. Calibration is assessed using the Brier score and Brier Skill Score (BSS). ΔBrier denotes the paired difference (DFS – Baseline); negative values indicate improved calibration with DFS.

| Horizon | Model | Prevalence | Null Brier | Brier (Base) | Brier (DFS) | BSS (Base / DFS) | ΔBrier (95% CI) |
| --- | --- | --- | --- | --- | --- | --- | --- |
| 30-day | LR | 0.1637 | 0.1369 | 0.2159 | 0.2165 | −0.577 / −0.582 | +0.0006 (−0.0003, +0.0015) |
| 30-day | LGBM | 0.1637 | 0.1369 | 0.1596 | 0.1572 | −0.166 / −0.148 | −0.0025 (−0.0029, −0.0020) |
| 30-day | XGB | 0.1637 | 0.1369 | 0.1612 | 0.1587 | −0.178 / −0.159 | −0.0025 (−0.0030, −0.0020) |
| 30-day | MLP | 0.1637 | 0.1369 | 0.1664 | 0.1689 | −0.215 / −0.234 | +0.0025 (+0.0019, +0.0032) |
| 60-day | LR | 0.2287 | 0.1764 | 0.2161 | 0.2159 | −0.225 / −0.224 | −0.0001 (−0.0009, +0.0006) |
| 60-day | LGBM | 0.2287 | 0.1764 | 0.1729 | 0.1701 | +0.020 / +0.035 | −0.0027 (−0.0032, −0.0022) |
| 60-day | XGB | 0.2287 | 0.1764 | 0.1759 | 0.1724 | +0.003 / +0.023 | −0.0035 (−0.0039, −0.0031) |
| 60-day | MLP | 0.2287 | 0.1764 | 0.1811 | 0.1976 | −0.027 / −0.120 | +0.0165 (+0.0159, +0.0171) |
| 90-day | LR | 0.2682 | 0.1963 | 0.2159 | 0.2202 | −0.100 / −0.122 | +0.0043 (+0.0034, +0.0051) |
| 90-day | LGBM | 0.2682 | 0.1963 | 0.1800 | 0.1765 | +0.083 / +0.101 | −0.0035 (−0.0039, −0.0030) |
| 90-day | XGB | 0.2682 | 0.1963 | 0.1835 | 0.1797 | +0.065 / +0.085 | −0.0038 (−0.0042, −0.0034) |
| 90-day | MLP | 0.2682 | 0.1963 | 0.1924 | 0.2020 | +0.020 / −0.029 | +0.0096 (+0.0089, +0.0103) |

Logistic regression exhibited minimal or negative calibration changes with DFS. At 30 and 60 days, Brier estimates were small and their confidence intervals included zero, indicating no clear calibration benefit. At 90 days, DFS was associated with worsened calibration for logistic regression, with the Brier score increasing from 0.2159 to 0.2202 (Brier +0.0043, 95% CI +0.0034 to +0.0051) and a corresponding decrease in Brier Skill Score.

We additionally report two-sided bootstrap p-values in Table 4. Across model families, the calibration results mirror the discrimination and operating-point analyses: DFS consistently improved calibration for boosted tree models, while offering little or no benefit for linear models.

Results for additional model families (XGBoost and multilayer perceptron), reported in the Supplementary Appendix, followed the same qualitative pattern: DFS reliably improved boosted tree models, while linear and neural models exhibited minimal or negative changes. Taken together, these findings demonstrate that the benefit of automated feature engineering is model-class dependent, with the largest and most consistent gains observed for tree-based learners across all readmission horizons.

## 4. Discussion

In this study, we evaluated whether automated feature engineering via Deep Feature Synthesis (DFS) improves the clinical applicability of heart failure (HF) readmission prediction from longitudinal EHR data. Across a large, real-world cohort spanning 15 years, we found that DFS provides consistent and meaningful benefits for gradient-boosted tree models, while offering little or no benefit for linear models. These gains extended beyond discrimination to include calibration and deployment-relevant operating characteristics, highlighting the model-class dependence of automated feature engineering in clinical prediction.

### 4.1. Principal Findings

DFS-enhanced features improved discrimination and precision–recall performance for LightGBM across all prediction horizons (30, 60, and 90 days), with AU-ROC and AUPRC gains that were consistent in magnitude and direction. At sensitivity-targeted operating points, DFS further increased specificity and positive predictive value, translating into a lower false-positive workload for a fixed level of case capture. Importantly, DFS also improved probability calibration for LightGBM at all horizons, with paired reductions in Brier score whose confidence intervals did not cross zero.

In contrast, logistic regression exhibited neutral or negative changes in discrimination, operating characteristics, and calibration when trained on DFS-enhanced features. These results indicate that the benefits of automated feature engineering are not universal, but instead depend strongly on the inductive biases and representational capacity of the down-stream model.

### 4.2. Why DFS Benefits Tree-Based Models but Not Linear Models

The observed model-class dependence is consistent with how different learners exploit expanded feature spaces. DFS generates a large number of aggregated and transformed features that encode temporal patterns, recurrence, and non-linear interactions across EHR domains (Kanter and Veeramachaneni, 2015; Fine et al., 2024). Gradient-boosted tree models are better suited as they can select informative features, model non-linear effects, and capture higher-order interactions without requiring explicit specification (Xu et al., 2014; Zhang et al., 2019).

By contrast, linear models rely on additive combinations of features and are sensitive to collinearity and noise introduced by feature expansion. In this context, DFS likely increases variance without improving the expressive power of the linear decision boundary, leading to degraded performance and calibration. This finding underscores that automated feature engineering should not be viewed as a drop-in improvement, but rather as a technique whose utility depends on alignment with the chosen model family.

### 4.3. Implications for Clinical Deployment

A key contribution of this work is demonstrating that DFS can improve deployment-relevant trade-offs, not just aggregate discrimination metrics. At matched sensitivity, DFS-enhanced LightGBM models reduced the number of false-positive alerts per true readmission, a metric directly tied to clinician workload and alert fatigue. Improvements in calibration further support the use of DFS-enhanced models for risk stratification and threshold-based decision-making, where reliable probability estimates are essential.

These findings suggest that automated feature engineering may offer a practical middle ground between manually curated feature sets and end-to-end representation learning. DFS can systematically incorporate longitudinal structure while remaining compatible with established, interpretable model families that are commonly deployed in healthcare settings.

### 4.4. Relation to Prior Work

Prior studies of HF readmission prediction have reported modest discrimination and limited calibration, even when using machine learning methods Yu and Son (2024); Van Grootven et al. (2021); Fine et al. (2024). Our results extend this literature by isolating the effect of feature construction from model choice and by evaluating performance across multiple horizons and operating regimes. While previous work has emphasized model architecture, our findings highlight feature engineering as a critical—and model-dependent—determinant of clinical utility (Abdul-Samad et al., 2024; Frizzell et al., 2017).

### 4.5. Limitations

This study has several limitations. First, data were drawn from a single urban safety-net health system, which may limit generalizability to other populations or care settings. Second, although DFS systematically expands the feature space, we constrained feature depth and applied filtering to manage computational cost and leakage risk; deeper or alternative configurations may yield different results. Third, each hospitalization was treated as an independent index event, which does not explicitly model within-patient temporal dependence across admissions. Finally, while we evaluated multiple model families, additional architectures and representation learning approaches were not explored.

### 4.6 Future Directions

Future work should examine the generalizability of these findings across institutions and patient populations, as well as the interaction between DFS and other model classes, including survival models and time-to-event frameworks. Further investigation into feature selection, pruning, and interpretability for DFS-generated features may also enhance clinical trust and adoption. Finally, prospective evaluation is needed to assess whether the observed improvements translate into better clinical workflows and patient outcomes.

### 4.7. Conclusion

Automated feature engineering via DFS consistently improved discrimination, calibration, and deployment-relevant operating characteristics for gradient-boosted tree models, while offering no benefit for linear models. These results demonstrate that the value of automated feature engineering is model-class dependent and that, when paired with appropriate learners, DFS can meaningfully enhance the clinical applicability of readmission risk prediction.

## Data Availability

All data produced in the present study are available upon reasonable request to the authors

## Appendix A.

**Figure 1.**
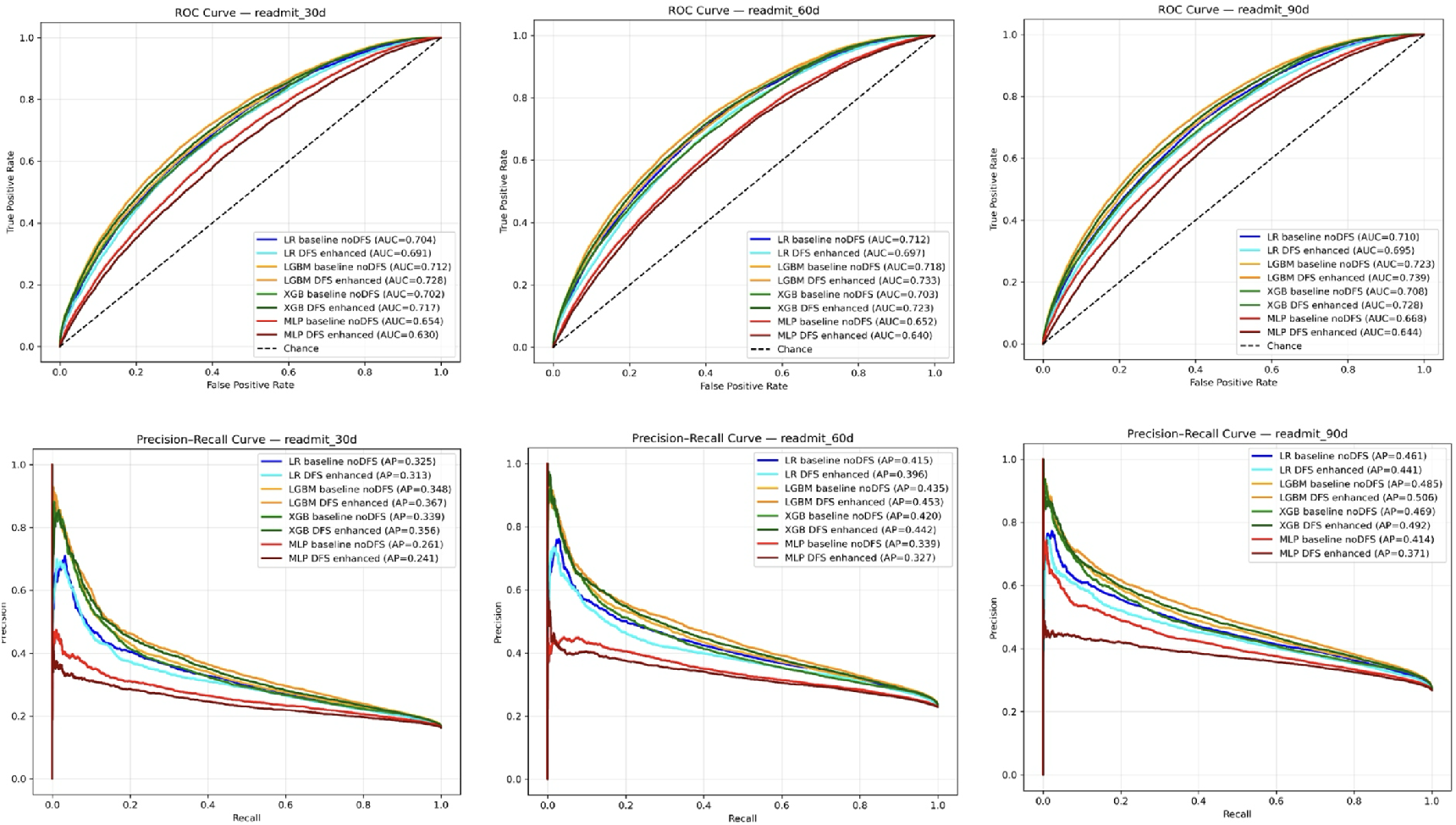
Receiver operating characteristic (ROC) and precision − recall (PR) curves for 30-, 60-, and 90-day readmission prediction.

## Footnotes

1 https://github.com/AnotherSamWilson/miceforest

2 Additional baseline clinical characteristics are provided in Supplementary Table S1.

